# Resetting public adherence: iterative strategies to counteract pandemic fatigue

**DOI:** 10.1101/2025.01.15.25320470

**Authors:** Albano Rikani, Laura Di Domenico, Chiara E. Sabbatini, Victor Navarro, Leo Ferres, Jocelyn Raude, Vittoria Colizza

## Abstract

Non-pharmaceutical interventions (NPIs) are essential for controlling infectious diseases during pre-vaccine periods, yet their success hinges on sustained public adherence. This study investigates adherence dynamics to tiered restriction systems implemented during COVID-19 in six geographical regions across Europe, North America, Africa, and South America. Using daily mobility data and linear-mixed models, we assessed three types of fatigue: overall fatigue (linked to cumulative time under restrictions), tier fatigue (linked to time spent under a specific tier), and iteration fatigue (linked to repeated implementation of the same tier). Tier fatigue caused the most rapid adherence loss, producing effects within days that overall fatigue required months of restrictions to achieve. Iterative application of shorter NPIs, interspersed with temporary relaxation, helped reset adherence, mitigating fatigue and sometimes even improving compliance. Psychological relief and a sense of regained autonomy during relaxation periods may renew public willingness to comply when restrictions are reintroduced. These findings emphasize the dual benefits of short, strategic NPIs for epidemic control and public resilience, offering actionable insights for designing more sustainable pandemic interventions.

## INTRODUCTION

The implementation of non-pharmaceutical interventions (NPIs) has served as a cornerstone strategy to mitigate the spread of infectious diseases during the critical periods before vaccines become widely available^1– 3^. In the context of the COVID-19 pandemic, NPIs were massively implemented^4,5^. Lockdowns, mobility restrictions, curfews, tiered systems and combinations thereof were critical in controlling disease transmission till summer 2021^6–9^. However, their prolonged application, coupled with high economic and social costs^10–12^, highlighted the need for strategic approaches to ensure adherence and effectiveness^13–16^. As emerging threats such as H5N1 loom^17^ and advancements in vaccine technologies like mRNA shorten vaccine deployment timelines^18^, it remains crucial to explore how pandemics can be effectively managed during the interim months before mass immunization.

The COVID-19 pandemic offers a valuable case study for understanding how human behavioral responses influence the success of NPIs^19–21^. During this pandemic, adherence to NPIs, commonly measured by the change of mobility over time, varied significantly, shaped by the overall duration of restrictions and the stringency of specific measures^22,23^. A growing body of evidence suggests that pandemic fatigue—a phenomenon characterized by declining adherence to restrictions over time^24,25^—can substantially undermine the effectiveness of interventions^13,26^. Moreover, modeling studies have shown that incorporating behavioral dynamics, such as reduced adherence in response to improving epidemic conditions, can lead to the emergence of multiple epidemic waves^19,27^.

Two key mechanisms driving this fatigue have been identified. The first, *overall fatigue*, reflects the cumulative time individuals spend under restrictions and has been observed globally^22^. The second, *tier fatigue*, relates to the stringency and duration of specific measures and has been extensively studied in the context of Italy’s tiered restriction system^23^. Evidence from Italy suggests that tier fatigue emerges rapidly, with adherence declining significantly within days under highly stringent measures. Beyond these mechanisms, another potential driver of fatigue may stem from the frequency of tier implementation, here termed *iteration fatigue*. This phenomenon may manifest as either growing leniency in adherence over successive iterations or, conversely, as a periodic renewal of public compliance as individuals re-engage with newly iterated measures. While the available evidence provides a foundation, gaps remain in understanding how fatigue operates across different geographical contexts, the extent to which adherence depends on the frequency of tier implementation, and the ways in which these mechanisms interact to shape public behavior. Can NPIs be strategized in such a way as to mitigate pandemic fatigue?

This study explores these gaps by analyzing the interplay between three types of pandemic fatigue—overall, tier, and iteration fatigue—across six diverse regions spanning Europe, Africa, and the Americas. Using COVID-19 as a case study, we investigate how these fatigue mechanisms impacted adherence to tiered restriction systems applied at different spatial scales and across different geographical contexts. By systematically assessing adherence dynamics, our findings aim to inform strategies for designing and implementing NPIs during future pandemics. This approach seeks to enhance early disease control efforts and ensure a smoother transition to vaccine-mediated mitigation.

## METHODS

Our study examines six geographical regions that implemented tiered restriction systems during the COVID-19 pandemic: Chile, Scotland, Ontario, California, Italy and South Africa. The characteristics of these systems varied across regions, including the number of tiers, their classification terminology, the specific restrictions assigned to each tier, and the geographical scale of implementation. For ease of presentation, we recoded the tiers for each system numbering them from 1 to 6, with increasing stringency of restrictions (Supplementary Data, **Table S1**), thus preserving the original number of tiers and their relative stringency. The administrative level at which the tiers were applied also differed, ranging from the municipal to the provincial or county level, referred to as the “area of implementation.” The mobility data we used is of the same or higher spatial resolution as the area of implementation of these tiers.

## Data

### Non-pharmaceutical interventions

We collected data on the timeline of tiered restrictions applied in each area within the studied regions (Supplementary data, **Figure S1**). The source for each dataset is reported in the Supplementary Data (**Table S2**).

In Chile, a five-tier system was implemented from July 25, 2020, to April 13, 2022, at the municipality or sub-municipality level. Data on the history of tiers applied, starting from July 28, 2020, was obtained from the Chilean Ministry of Health. However, due to significant changes to the tier system on September 30, 2021, we focused our study on the period from July 28, 2020, to September 30, 2021.

A five-tier system was also used by the Scottish government from November 2, 2020 to August 9, 2021. We retrieved data on the tier implementation timeline from the UK’s archived repository. In Scotland, restrictions were primarily applied at the council area level, with a few exceptions for small islands, where a finer spatial scale was used. In such cases, we assigned the tier of the most populated sub-area to the entire council (e.g., Argyll and Bute). Additionally, from January to April 2021 Scotland underwent a national lockdown. Since the national lockdown had a higher level of stringency than tier five, we categorized this phase as tier six.

In Canada, the province of Ontario implemented the COVID-19 Response Framework “Keeping Ontario Safe and Open” from November 7, 2020, to May 1, 2021. This five-tier system was applied at the Public Health Unit (PHU) level, which are administrative divisions responsible for managing health services. From December 26, 2020, to mid-February 2021, Ontario underwent a province-wide shutdown. We used the same approach as for Scotland’s national lockdown, incorporating the shutdown as a sixth tier. The timeline of applied tiers and the shutdown was obtained from the Ontario government.

In the United States, the state of California adopted the “Blueprint for a Safer Economy” plan on August 31, 2020, which remained in effect until June 14, 2021. This system included four tiers applied at the county level. On December 5, 2020, the state government of California introduced a regional stay-at-home order, which was more stringent than tier four. As a result, we classified the stay-at-home phase as tier five. Data on the timeline of tier assignments were obtained from the California Department of Public Health.

The Italian government introduced a four-tier restriction system on November 6, 2020, which was applied at the regional level, including the autonomous provinces of Bolzano and Trento. This system remained in place until March 31, 2022. Data on the timeline of tier implementation was obtained from the repository of the Italian Civil Protection Department.

The government of South Africa established a five-tier alert system following a national lockdown at the end of March 2020. This alert system was implemented nationwide from March 27, 2020, to April 4, 2022. Data on the applied tiers were collected from official government reports.

We provide an overview of the main restrictions associated with each tiered system in the Supplementary Data (**Table S3**). During the implementation period, the tiered systems underwent adjustments in terms of the specific restrictions linked to each tier. The key changes for each system are also detailed in the Supplementary Data.

### Mobility data

We used two sources of mobility data, focusing on two key metrics that reflect behavioral changes compared to pre-pandemic levels for each source: changes in residential time and visits to workplaces.

The first source was the Google Mobility Reports^28^, which tracked daily mobility changes from February 15, 2020, to October 15, 2022. These reports provide relative changes in time spent at residences and in the total number of visits to workplaces, calculated against baseline values. The baselines are the median values obtained from the pre-pandemic period spanning January 3 to February 6, 2020. As mobility data from Google Mobility Reports was not available at the municipality level in Chile, the second source of mobility data was from the mobile phone operator Telefónica Movistar in Chile, covering the period from March 2020 to December 2021. This dataset consists of eXtended Detail Records (XDR)^29^, which collect information on the time and tower location of connections for each device. We defined two metrics to capture residential time and trips to workplaces, respectively (see Supplementary Methods). To align this data with the Google Mobility Reports, we used baseline weekday-specific values calculated as the mean from the longest available pre-pandemic period, March 2–15, 2020.

Both data sources cover the entire period of implementation of the tiered restrictions. Therefore, our periods of study correspond to the entire duration of the tiered systems in each geographical region, unless otherwise stated. Specifically, for South Africa it goes from March 27, 2020, to April 4, 2022. For Italy, the period of study starts on November 6, 2020 and ends on March 31, 2022. For California we cover the period from August 31, 2020 to June 14, 2021, while for Ontario we use the period from November 7, 2020, to May 1, 2021. For Scotland we used the period that goes from November 2, 2020 to August 9, 2021. Finally, for Chile we used a reduced period of study from July 28, 2020, to September 30, 2021. In some regions, the spatial scale at which NPIs were applied differed from the spatial resolution of mobility data. When the mobility data had a higher resolution than the NPIs, we downscaled the NPIs to match the mobility data, as was done for Ontario and South Africa. For South Africa, restrictions were applied nationwide, but mobility data was available at the province level. In contrast, in Chile, some restrictions were applied at the sub-municipality level, while our mobility data was aggregated at the municipality level. As a result, we excluded 36 municipalities (out of 346) from our analysis (see Supplementary Data).

## Models

We assessed the impact of three different types of pandemic fatigue on adherence to non-pharmaceutical interventions (NPIs) that limited mobility:

- *Overall fatigue*: This refers to the cumulative time spent under restrictions, regardless of the stringency level. It is measured as the total number of days (*T*_*a,d*_) that an area *a* has spent under restrictions by a given day *d* since the tiered system was implemented.
- *Tier fatigue*: This refers to the time spent in each specific tier during each implementation. We measured it as the number of days 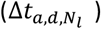 that an area *a* on day *d* has spent in tier *l* during its *N*-th implementation.
- *Iteration fatigue*: This refers to the number of times (*N*_*a,l,d*_) that a given tier *l* has been implemented in an area *a* by day *d*.

Our analysis used linear mixed models (LMMs). We first defined a set of three baseline LMMs, each corresponding to one of the three types of fatigue.

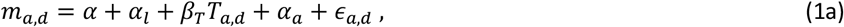

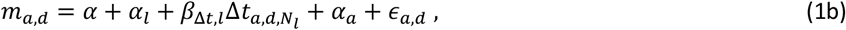

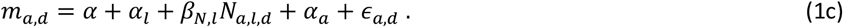

Model 1 of Eq.1a incorporates overall fatigue; Model 2 of Eq.1b incorporates tier fatigue; and Model 3 of Eq.1c incorporates iteration fatigue. Here, *m*_*a,d*_ represents the daily change in mobility in area *a* (e.g., municipalities in Chile, counties in California, etc.) on day *d*. We controlled for area-specific factors using area-specific intercepts *α*_*a*_, which were modeled as random effects. Each model also includes a global intercept *α* and a tier-specific intercept (fixed effect) *α*_*l*_. The error term is denoted by *ϵ*. Notably, 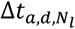 is an area- and tier-specific day counter, which resets to zero at the end of each implementation *N*_*l*_ of tier *l* in area *a*. A schematic representation of the three types of fatigue can be found in the Supplementary Methods (**Figure S2**).

Since our analysis period spans different seasons, the change in mobility, particularly in terms of residential time, may reflect seasonal patterns. Additionally, seasonality effects have been observed in SARS-CoV-2 transmission^30,31^, which could influence behavior through changes in risk perception^32^. To account for these effects, we introduced a seasonality variable *S* modeled using a sinusoidal function with its peak on January 22.

This date roughly corresponds to the average peak of seasonality of transmission observed in the French regions^33,34^:

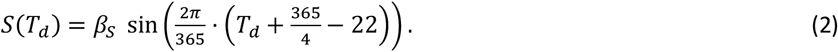

The variable *T*_*d*_ is a year-specific counter, computed as the number of days from the beginning of the year up to day *d*. For countries in the Southern Hemisphere (i.e., Chile and South Africa), we adjusted *S* by adding *π* to the sinusoidal function in Eq.2. This shift aligns the peak of the adjusted *S* function with the day when *S* reaches its minimum value.

Using Eqs. 1a-1c and Eq. 2, we defined five additional LMMs of increasing complexity, incorporating multiple types of fatigue simultaneously and controlling for the seasonality effect:

- Model 4 includes both overall and tier fatigue.
- Model 5 includes both overall and iteration fatigue.
- Model 6 includes both tier and iteration fatigue.
- Model 7 includes all three types of fatigue.
- Model 8 builds upon Model 7 by adding the seasonality term.

Our most comprehensive specification is therefore Model 8, which assumes the following form:

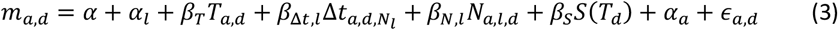

Lastly, we controlled for the effect of the population’s perceived epidemiological risk on adherence. To do this, we defined Models 9 and 10 by extending Model 8 to include the incidence of new cases and its interaction with the time spent under restrictions, respectively (see Supplementary Methods). However, the incidence datasets do not cover the entire period of study and all areas of each geographical region, ultimately limiting the ability of Models 9 and 10 to fully investigate the three types of fatigue. Therefore, we used these models solely for sensitivity analysis. The full expression of each model is reported in the Supplementary Materials.

We ran each LMM separately for each of the six geographical regions in our study, obtaining estimates of ***α*** *=* {*α, α*_*l*_} and ***β*** *=* {*β*_*T*_, *β*_Δ*t,l*_, *β*_*N,l*_, *β*_*S*_}. We restricted our analysis to the tiers that occurred at least three times and we excluded those that by definition did not affect mobility (i.e., tier 1 in Italy) (Supplementary methods, **Figure S3**). We also removed bank holidays from the analysis. Importantly, if the implementation of the tiers is coordinated among the areas the effects of the iteration and the overall fatigue might not be distinguishable. Indeed, the coordination could lead to a high collinearity between the number of days spent under restrictions and the number of iterations. We evaluated such collinearity for each model using the Variance Inflation Factor (VIF). We calculated both the Akaike information criterion (AIC) and the marginal R^2^ for each model and used these quantities for selecting our preferred model specification.

## RESULTS

We found evidence of waning adherence due to tier fatigue across all regions and models, for both mobility metrics and for most tiers (**Figure 1** and Supplementary Results, **Figures S4** and **S5, Tables S4–S15**). Similarly, our results indicate that iteration fatigue led to significant declines in adherence across all models in Italy and South Africa for both mobility metrics, and in Chile and California for residential time (**Figure 1** and Supplementary Results, **Figures S6** and **S7, Tables S4–S15**). Additionally, we observed waning adherence due to overall fatigue in all models for Scotland and Chile (residential time), as well as for Italy (workplace trips) (**Figure 1** and Supplementary Results, **Figure S8, Tables S4–S15**). Notably, Italy (workplace trips) and Chile (residential time) exhibited significant waning adherence across all three types of fatigue.

**Figure 1:**
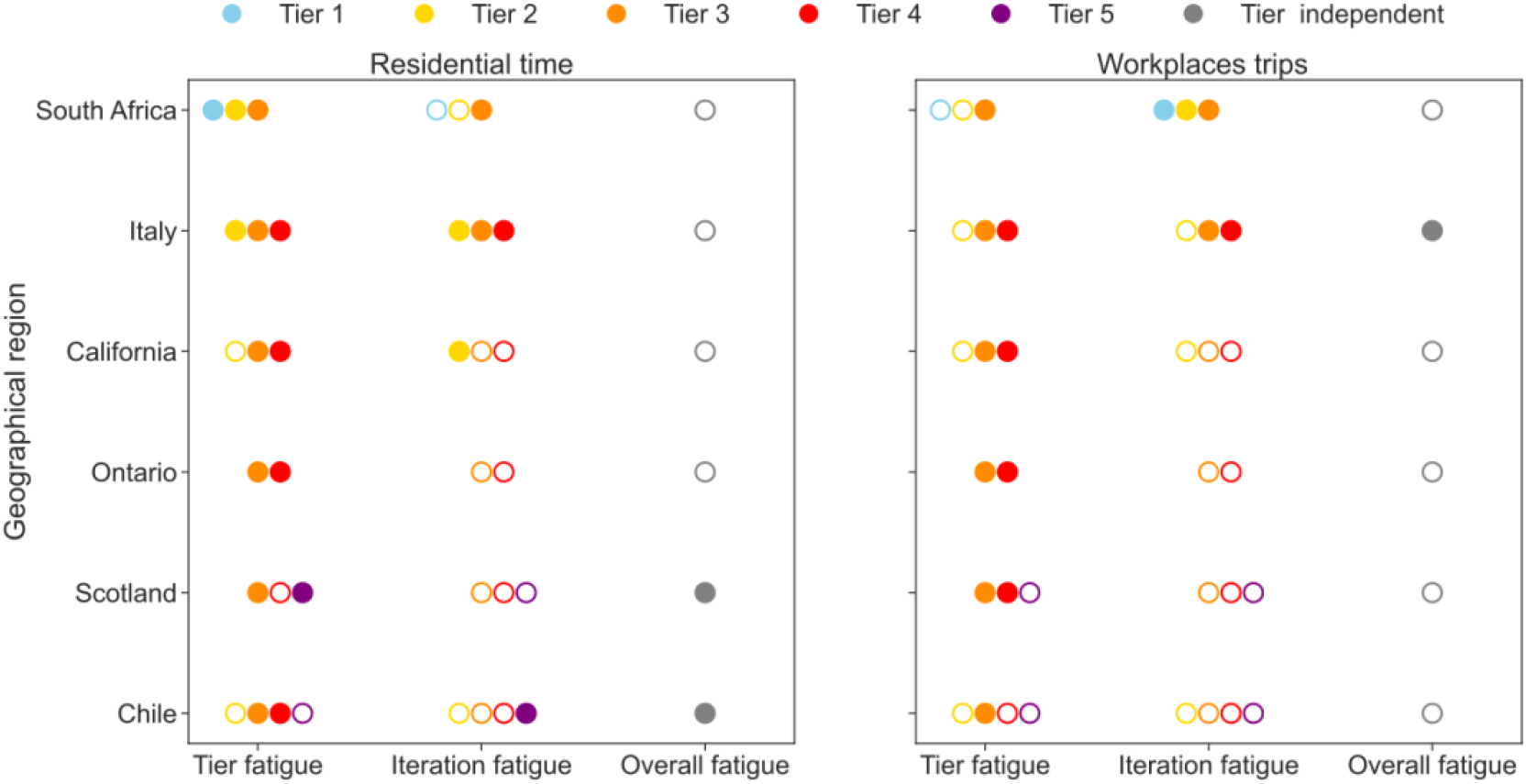
Type of loss of adherence observed in each geographical region for each mobility metric. Each point represents one type of fatigue and tier of restrictions. The overall fatigue is tier-independent. Filled dots represent results that are statistically significant and of the same sign across all models. Dots are otherwise drawn empty whereas missing dots represent tiers that were not included in the analysis or were not part of the tiered system in place in the region. Results obtained from using the residential time and trips to workplaces data are reported in the left-hand side and right-hand side panel respectively.

According to the marginal R2, each type of fatigue explains a similar portion of the variability in our data (**Figure 2**, Models 1–3). The explanatory power increases when we simultaneously include more than one type of fatigue, reaching a maximum when all three types of fatigue and the seasonality effect are included (**Figure 2**, Models 4– 8). Model 8 also performs the best in terms of the AIC (see Supplementary Results, **Figure S9**). This emphasizes the importance of simultaneously accounting for all three types of fatigue and controlling for seasonality effects. Since we observe similar results between the two mobility metrics, and because seasonality effects are easier to interpret for changes in residential time rather than trips to workplaces, we will focus on the residential time data and Model 8 in the following. The results for the remaining models, as well as for changes in workplace trips, are provided in the Supplementary Materials.

**Figure 2:**
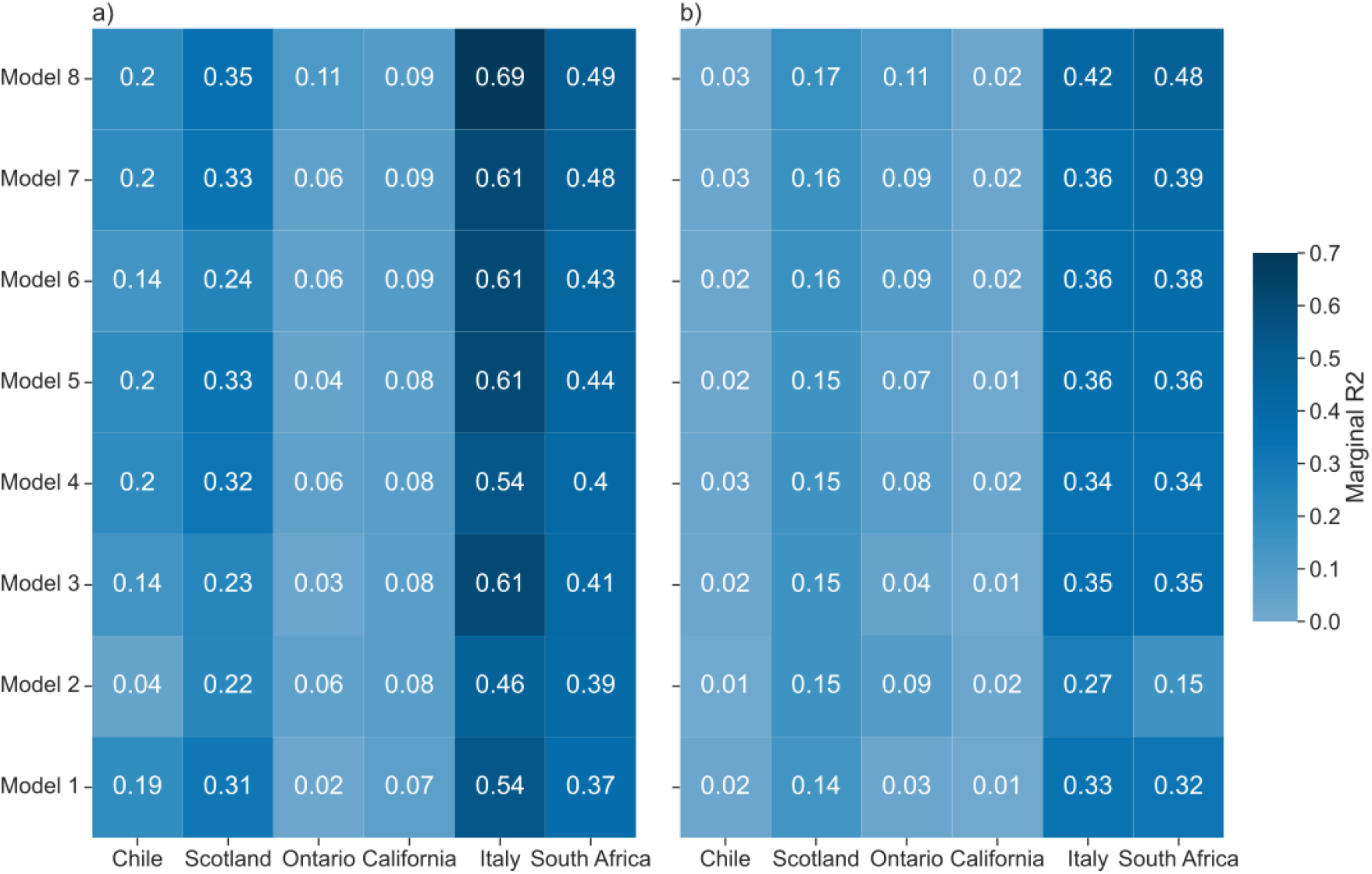
Marginal R2 for each model and region, for the residential time (a) and trips to workplaces (b).

Starting with overall fatigue, we found that most regions experienced a loss of adherence as the number of days under restrictions increased (**Figure 3a**). The strongest effect was observed in Chile, while it was smaller and comparable across Scotland, California, and Italy. In contrast, we did not find evidence of overall fatigue in Ontario and South Africa. Interestingly, in South Africa, results showed a strong positive effect, suggesting that compliance with NPIs increased over time. However, this result was driven by high collinearity between overall and iteration fatigue. In models that included overall fatigue but not iteration fatigue (i.e., Models 1 and 4), we observed a loss of adherence due to overall fatigue (see Supplementary Material, Figure S8). To evaluate the strength of the collinearity, we calculated the variance inflation factor (VIF). For South Africa, the VIF values were significantly higher than the critical threshold of 10 (see Supplementary Results, **Table S2**), while the remaining regions showed smaller and non-critical VIF values.

**Figure 3:**
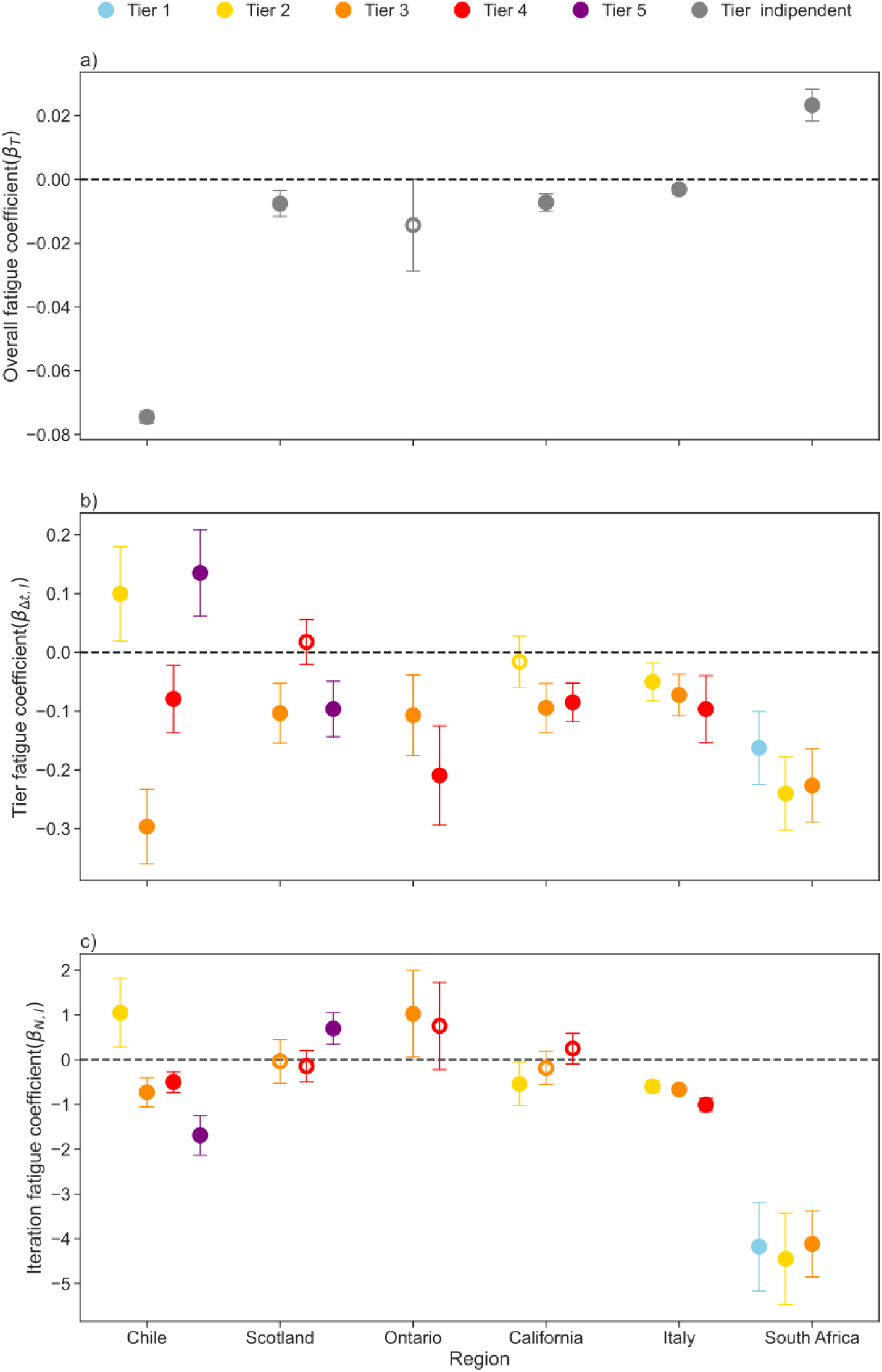
Estimates of the overall fatigue (a), tier fatigue (b) and iteration fatigue (c) for each region using data on change in residential time and Model 8. Regions are sorted in ascending order of the geographical area at which NPIs were applied, from municipality (Chile) to country (South Africa). Whiskers show the values at 95% confidence interval. Empty circles represent estimates statistically not significant at 95% confidence level.

Regarding tier fatigue, we found that it led to a loss of adherence across all regions and for almost every tier (**Figure 3b** and Supplementary Results, **Figure S4**). Moreover, in Italy and Ontario more stringent was the tier, greater was the observed loss of adherence. Notably, the strongest effects of loss of adherence were seen in regions where no overall fatigue was detected, specifically South Africa and Ontario. Finally, iteration fatigue led to waning adherence in Chile, California, and Italy (**Figure 3c**). In Chile, we found a significant loss of adherence under the three most stringent tiers, with the largest losses observed under the most stringent tier. In Italy, adherence declined for all three tiers, with stricter tiers showing stronger effects. Scotland and Ontario were the only regions where iteration fatigue did not result in a loss of adherence.

Our results remained robust when expanding Model 8 to control for risk adaptation and habituation effects (Supplementary Results, **Figures S11-S15**). While the inclusion of these factors improved the model’s explanatory power and was preferred in terms of AIC (Supplementary Results, **Figures S16** and **S17**), their impact on fatigue estimates was small and negligible.

So far, we have presented the results for each type of fatigue separately. However, understanding the best implementation strategy for tiered systems requires comparing the effects of the different types of fatigue. To this end, we characterized each type of fatigue by the number of days or iterations needed to produce a given level of adherence loss. We then translated the iterations into days based on the observed median duration of each tier in each region. For comparison, we focused on regions and mobility metrics that showed significant effects of all three types of fatigue. Here, we present the case of Italy using the residential time mobility data. Results for California, and Chile (residential time mobility data) and Italy (workplace mobility data) are provided in the Supplementary Results (**Figures S18–S20**).

For simplicity, we set the level of change in residential time to 2% and examined how quickly each type of fatigue reached this threshold. In Italy, iteration fatigue would lead to a 2% reduction in residential time after two iterations of tier 4 or three iterations of tier 3 (**Figure 4a**). The median duration of tier 4 was 25 days, while tier 3 lasted 20 days (**Figure 4d**). Thus, a sequence of tier 4 (25 days), followed by tier 3 (20 days), and then another tier 4 (25 days) would take 70 days to achieve a 2% reduction in adherence. In contrast, overall fatigue produced a much weaker effect, leading to less than a 0.1% reduction in residential time after 70 days under restrictions. Even after 500 days of restrictions—approximately the total time the country spent under restrictions (**Figure 4c**)—the reduction in residential time remained below 2% (**Figure 4b** inset).

**Figure 4:**
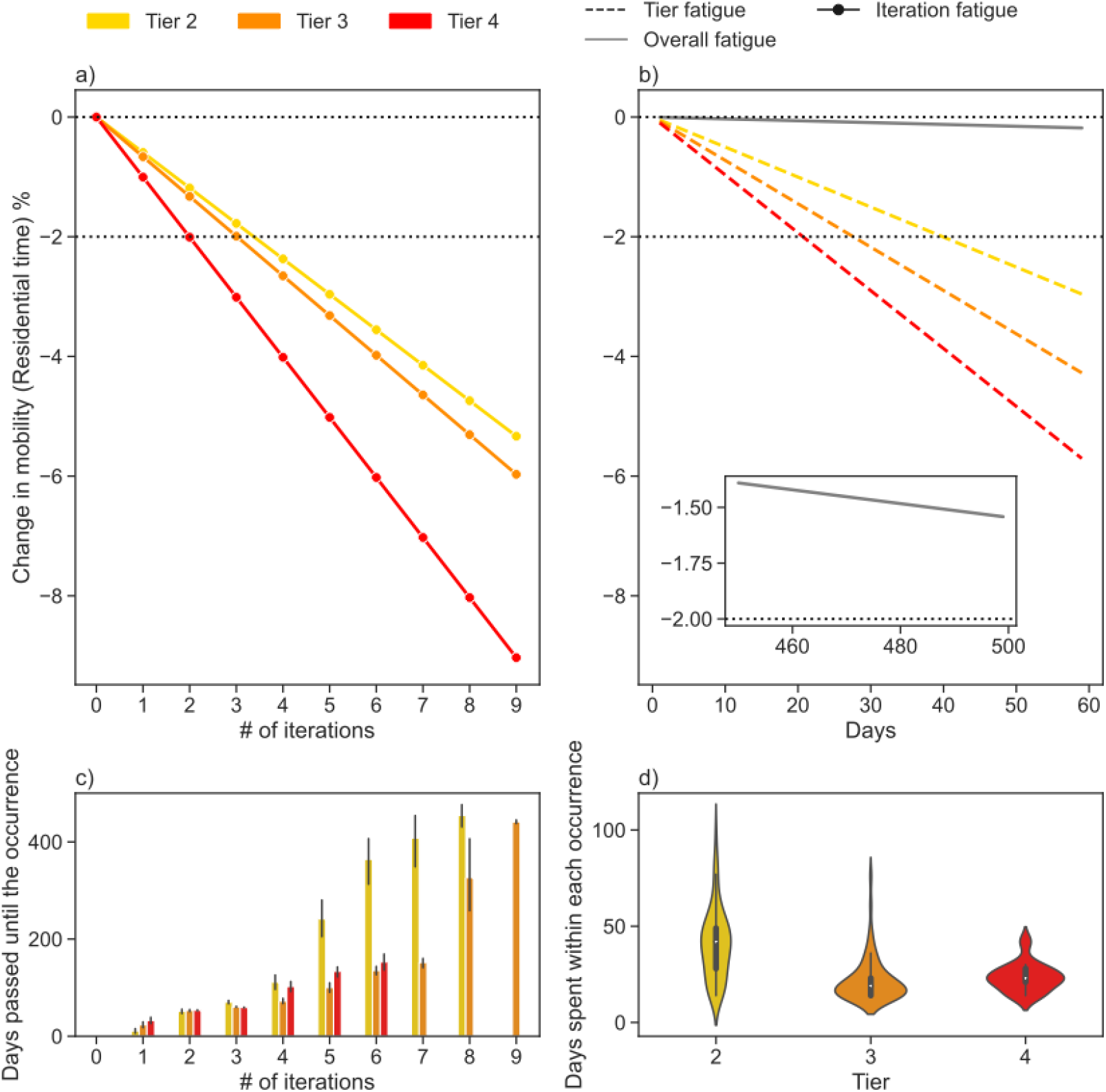
Estimated change in residential time in Italy due to the three types of fatigue and observed duration and iterations of the tiers. Panel (a) represents the effect from the iteration fatigue whereas panel (b) shows the effect due to the overall fatigue (grey line and inset) and the tier fatigue (b, colored lines). Panel (c) represents, for each tier, the average number of days passed from the initial implementation of the tiered system to the n-th iteration of the tier. Panel (d) shows for each tier the distribution of the duration of implementation for implementations that lasted longer than two weeks. While panels (a) and (b) represent estimates according to our modeling results, panels (c) and (d) represent historical data on the implemented strategy in Italy. The dashed line at -2% represents a specific level of change in mobility that we choose for discussing the results.

Tier fatigue, on the other hand, had a stronger effect, leading to a 2% reduction in residential time after approximately 40 days of tier 2 (**Figure 4b**). This effect was even stronger than iteration fatigue, which required more than 120 days (or three iterations of the same tier) to achieve the same change in residential time. Notably, the impact of tier fatigue was particularly strong in California (**Figure S18**), whereas it was negligible in Chile. In Chile, with NPIs applied at the municipality level, overall fatigue emerged as a dominant factor, with effects comparable to those of tier and iteration fatigue (**Figure S19**).

## DISCUSSION

NPIs that limit interpersonal contact were essential in mitigating the spread of COVID-19 while waiting for mass vaccination^6^. However, the individual cost of these measures often led to adherence fatigue, a phenomenon that was observed in multiple regions and contexts during the pandemic^22,23,26^. Understanding when and how this fatigue develops is essential for improving preparedness plans for future pandemics. In this study, we used data from six geographical regions that implemented tiered systems of interventions to explore the effects of three distinct types of fatigue— overall fatigue, tier fatigue, and iteration fatigue —on public adherence to NPIs.

Our findings showed that adherence waned significantly in response to tier and iteration fatigue, while the impact of overall fatigue was comparatively weaker. Specifically, overall fatigue required hundreds of days to produce adherence losses comparable to those observed after just ten days of tier fatigue or a few iterations of interventions. This stark contrast highlights the critical importance of the duration and frequency of interventions in shaping public compliance. Frequent, shorter interventions appear to be more effective in mitigating fatigue-related adherence loss than prolonged measures, emphasizing the need to account for these dynamics when designing NPIs.

Successive iterations can also reset adherence. Iteration fatigue was indeed generally absent in regions like Scotland, Ontario, and California, except for the most stringent tiers in Scotland (Tier 5) and Ontario (Tier 3) where adherence increased with successive iterations (similarly observed in Tier 2 in Chile as well). This may reflect a renewed compliance following breaks in restrictions, which provided psychological relief. Temporarily relaxing measures before reintroducing them could help reset public perceptions and expectations, provided such strategies are tailored to the local epidemiological conditions. Circuit-breakers emerge therefore as a valuable tool to manage iteration fatigue while maintaining control over the epidemic^13,35^. The increase in public adherence could also signal an evolution in public understanding of NPIs. Initial iterations of a tier may have been poorly understood or less favorably received, because of the novelty of the intervention, with adherence improving as the public became more familiar with the measures^36^. Clear, consistent, and transparent communication is therefore essential when implementing NPIs^37^. Enhanced communication and the consideration of circuit-breakers may also be important to counteract the stronger decline in compliance associated to stricter tiers, observed clearly in Italy^23^ and Ontario, and previously reported globally for high-cost interventions, compared to lower-cost measures like mask mandates^22^. Incremental adjustments to restrictions over time (Supplementary Data), to align measures with evolving practical realities, may have also contributed by easing compliance from one iteration to the next. However, these changes were typically minor and unlikely to fully account for the observed improvements.

Our study focuses on adherence to mobility restrictions and does not directly address the downstream epidemiological impacts of these behavioral changes. Previous research already linked tier fatigue to increased epidemic activity. For instance, a study in France associated tier fatigue with a rise in the reproduction number, driven by increased mobility and transmission during intervention periods^13^. The study also demonstrated that short, stringent lockdowns were more effective than prolonged, moderate measures in achieving better hospitalization outcomes, while causing similar levels of intermediate distress and infringement on individual freedoms. Similarly, research from the UK highlighted that planning limited duration periods of NPIs could effectively manage the epidemic while reducing societal impact^35^. Our findings further support prioritizing shorter restrictions, as they also reduce adherence loss. Adaptive strategies alternating between brief restrictions and periods of relaxation strike a balance between controlling viral transmission and addressing the behavioral and psychological dynamics of adherence. Such approaches minimize the strain on populations while maintaining the effectiveness of NPIs, fostering greater resilience during prolonged pandemics.

Nonetheless, some level of fatigue is inevitable. For instance, tiers often needed to remain in place for a minimum duration (e.g., two weeks in California, Chile, and Italy) to allow sufficient time for evaluation and potential adjustments. While necessary for stability and evaluation purposes, this approach imposes a baseline level of unavoidable tier fatigue that must be factored into intervention planning to optimize both adherence and effectiveness.

While fatigue-related mechanisms are central to our analysis, we also acknowledge that voluntary behavior changes influenced by risk perception may have played a role in adherence patterns. Previous studies highlighted a positive relationship between heightened risk perception and compliance with low-cost health protective behaviors, such as wearing masks and avoiding physical contact^38,39^. Although we lacked direct survey data on public risk perception at the resolution scale under study, we used epidemiological indicators, such as incidence rates, as proxies^32^. Our findings support the hypothesis of a risk adaptation mechanism, where higher levels of perceived risk (e.g., during periods of high incidence) are associated with stronger adherence to restrictions (Supplementary results, **Tables S16 - S27**)^40,41^. However, this effect did not substantially alter the observed impacts of fatigue, reinforcing previous findings that adherence decline cannot be fully explained by risk perception alone^22,23^. Another potential factor influencing adherence is risk habituation, a behavioral mechanism in which prolonged exposure to a health threat leads individuals to underestimate or neglect the risk, resulting in reduced compliance over time^40^. We tested for this hypothesis in sensitivity analyses and found that while risk habituation may contribute to adherence loss, the effects of tier and iteration fatigue remained robust. This suggests that fatigue-related mechanisms are likely the dominant drivers of adherence waning, rather than solely psychological desensitization to risk.

Our study is subject to a set of limitations that should be considered when interpreting the findings. First, the analysis does not explicitly account for spatial correlations. While the spatial scale in most regions was large enough to minimize the impact of inter-area trips, this may be more relevant in regions like Chile and California, where restrictions were applied at finer scales. However, in most cases restrictions prohibited movement between areas, likely reducing possible spatial correlation effects. Second, the mobility data used may not fully represent the entire population. However, both Google mobility and Telefonica Movistar data provide reliable proxies for mobility trends and have high coverage of the populations studied^23,42,43^. Third, using two different mobility data sources—Google for most regions and Telefonica Movistar for Chile because of resolution needs—may introduce inconsistencies. However, aggregating Telefonica Movistar data to the regional level in Chile showed strong correlation with Google data^44^, and estimates for the overall fatigue at the regional level were comparable to those for the other geographical regions (Supplementary results, **Figure S22**). This suggests that observed differences are likely due to contextual factors rather than data inconsistencies. Lastly, our study is restricted to six geographical regions as those are the ones that implemented tiered NPIs, allowing us to study the multi-factorial aspects of adherence loss dynamics.

NPIs have proven to be a critical first-line response during pandemics^2,3^ and are likely to remain a key tool in managing future infectious disease threats. Our study provides a detailed understanding of the impact of fatigue on adherence to tiered interventions, emphasizing the need to account for tier and iteration fatigue in preparedness planning. Our findings highlight the importance of balancing stringency, duration, and frequency in designing NPIs. Strategic use of circuit-breakers and adaptive intervention planning can play a crucial role in maintaining adherence while ensuring the effectiveness of public health measures. Clear communication and public engagement are also key to fostering sustained compliance and mitigating the effects of fatigue. By integrating these insights into preparedness planning, policymakers can better navigate the challenges of public compliance and improve the management of future pandemics.

## Data Availability

Data from Google mobility reports are publicly available. Mobility data for Chile produced in the present study are available upon reasonable request to the authors.

https://www.google.com/covid19/mobility/

## ACKNOWLEDGMENTS

The authors thank Shweta Bansal for useful discussions on this study. The research was partially supported by: ANR grant DATAREDUX (ANR-19-CE46-0008-03) to LDD, CES, VC; EU Horizon 2020 grant MOOD (H2020-874850, publication cataloged as MOOD 124) to AR, CES, VC; EU Horizon Europe grant VERDI (101045989) to VC; EU Horizon Europe grant ESCAPE (101095619) to AR, VC; Telefónica R&D Chile and CISCO Chile to LF; FONDECYT Grant N°1221315 to LF; Lagrange project of the ISI Foundation funded by Fondazione CRT to LF. The contents of this publication are the sole responsibility of the authors and don’t necessarily reflect the views of the European Commission.

